# Biased and unbiased estimation of the average length of stay in intensive care units in the COVID-19 pandemic

**DOI:** 10.1101/2020.04.21.20073916

**Authors:** Nathanael Lapidus, Xianlong Zhou, Fabrice Carrat, Bruno Riou, Yan Zhao, Gilles Hejblum

## Abstract

**Background:** The average length of stay (LOS) in the intensive care unit (ICU_ALOS) is a helpful parameter summarizing critical bed occupancy. During the outbreak of a novel virus, estimating early a reliable ICU_ALOS estimate of infected patients is critical to accurately parameterize models examining mitigation and preparedness scenarios.

**Methods:** Two estimation methods of ICU ALOS were compared: the average LOS of already discharged patients at the date of estimation (DPE), and a standard parametric method used for analyzing time-to-event data which fits a given distribution to observed data and includes the censored stays of patients still treated in the ICU at the date of estimation (CPE). Methods were compared on a series of all COVID-19 consecutive cases (n=59) admitted in an ICU devoted to such patients. At the last follow-up date, 99 days after the first admission, all patients but one had been discharged. A simulation study investigated the generalizability of the methods’ patterns. CPE and DPE estimates were also compared to COVID-19 estimates reported to date.

**Findings:** LOS ≥ 30 days concerned 14 out of the 59 patients (24%), including 8 of the 21 deaths observed. Two months after the first admission, 38 (64%) patients had been discharged, with corresponding DPE and CPE estimates of ICU_ALOS (95%CI) at 13.0 days (10.4–15.6) and 23.1 days (18.1–29.7), respectively. Series’ true ICU_ALOS was greater than 21 days, well above reported estimates to date.

**Interpretation:** Discharges of short stays are more likely observed earlier during the course of an outbreak. Cautious unbiased ICU_ALOS estimates suggest parameterizing a higher burden of ICU bed occupancy than that adopted to date in COVID-19 forecasting models.

**Funding:** Support by the National Natural Science Foundation of China (81900097 to Dr. Zhou) and the Emergency Response Project of Hubei Science and Technology Department (2020FCA023 to Pr. Zhao).

## 1. Introduction

The spread of a novel coronavirus (SARS-CoV-2) has brought about a pandemic referred to as the COVID-19 pandemics.^1^ This pandemic has resulted in a worldwide crisis with unprecedented decisions of restrictive non-pharmacological mitigation interventions taken at local, regional, or national levels. A major aim of these measures is lessening as much as possible the daily number of new individuals requiring an admission in intensive care units (ICU) in order to be able to appropriately manage them in the healthcare system and sustain an appropriate management for the rest of the population.^2^ A fast inflow of new admissions in the ICU has critical consequences within a short time. For example, between March 19 and April 2 2020 in France, the number of ICU beds occupied by COVID-19 infected persons dramatically increased from 1002 to 6305,^3^ corresponding to an average daily increase of 14 % additional beds. Such a situation requires a massive and rapid increase of ICU facilities and the French Minister of Health announced on March 28 that the nationwide capacity had been increased from 5000 to 10,000 critical beds.^4^ The underlying mathematics are simple: an average unbalanced increase of 15% during 14 days implies that at day 14, the resulting occupancy would be that of day 0 multiplied by a factor 7.08 since 1.15^(14 days)^ = 7..08 The system is highly sensitive to a sustained unbalance: even an average increase as low as 2% during two weeks, a likely situation after outbreak peak, would nevertheless require increasing occupancy at day 14 by 32%.

The average length of stay (ALOS) in ICU is an important estimate relating to the stability of the healthcare system in terms of ICU bed occupancy, as illustrated by the following situation:

- Let us consider that the needs in ICU beds under study only concern patients infected by a new emerging agent, and let us assume that such a patient population would have an ICU ALOS equal to 10 days.
- Then, the daily probability of a bed discharge would be 1/ALOS = 1/10 = 0.10. This implies that every day, in average, 10% of the occupied beds in the ICU should concern the discharge of the corresponding patient.
- This implies in turn that the above calculated ALOS-dependent threshold of 10% of the capacities constitutes the equilibrium point of the system: whenever the rate of required admissions is exactly 10%, the daily number of new admissions is likely perfectly balanced with the daily number of discharges, and therefore the total number of beds occupied remains stable from one day to another. However, whenever the rate of required admissions would exceed the 10% ALOS-dependent threshold, the global number of beds occupied or required would increase while conversely, whenever this rate would be below the 10% ALOS-dependent threshold, the beds occupied or required would decrease.

This example demonstrates that estimating the ICU ALOS of a population infected by an emergent virus constitutes a very critical information to modelers and decision makers for guiding adaptations of the local capacities in the context of the outbreak. Such an estimate is expected to be provided as soon as possible. However, when examining the situation within a short delay after the beginning of the outbreak, only few cases are likely to be already discharged from the ICU. The patients still in ICU referred to as censored cases must be considered in any unbiased estimation relating to the length of stay (LOS). In this study, we present a detailed examination of the timeline of the whole cohort of consecutive COVID-19 patients admitted to a devoted ICU of the Zhongnan hospital of Wuhan University (ZHWU) in which we investigated the evolution of the ALOS estimation according to the accumulation of the cases, using two methods of estimation. Our results indicate that even considering a last follow-up date corresponding to the date when two thirds of the admitted patients would have been discharged, the ICU ALOS estimated with the biased method would be nearly half of that issued from the unbiased method. At the light of these investigations, the estimates relating to ICU LOS of COVID-19 cases that have been reported to date^5-8^ likely underestimate the real values. Such estimates being also used in forecasting models,^9-13^ the present study has practical implications for improving prediction scenarios to guide public decision.

## 2. Methods

### Ethics

This study was approved by the Medical Ethics Committee, ZHWU (Clinical Ethical Approval No.2020005). The informed consent was waived by the Medical Ethics Committee for emerging infectious disease.

### Setting

As in many locations, the organization of the ZHWU (Hubei province, Peoples’s Republic of China) for managing COVID-19 patients was subjected to several changes during the course of the COVID-19 outbreak. First, on December 30 2019, at a time when the outbreak emerged frankly, two initial ICU, one depending on emergency and the other from surgery, were reorganized for constituting a single entity of 31 beds devoted to the management of patients with COVID-19 requiring critical care. Second, on March 12 2020, at a time when the outbreak had declined, all COVID-19 ICU patients were transferred to another ICU in Leishenshan hospital, the largest newly-built facility for COVID-19 patients with 1,600 beds, while ICU admissions were reorganized for other pathologies than COVID-19 at ZHWU. Third, on April 15, Leishenshan hospital was definitively closed and patients initially admitted at ZHWU were retransferred to this hospital.

### Patients

All consecutive patients with a confirmed diagnosis of COVID-19 by PCR and initially admitted to the above-mentioned ICU of 31 beds at ZHWU from December 30 2019 to March 12 2020 (n = 57) were included in the study. Patients admitted to this ICU during this period also included 10 consecutive patients for which there was a radiological evidence of viral pneumonia^14^ while RT-PCR test of throat swabs had remained negative for several times, and these patients were also considered as eligible for the study. Last follow-up of patients was made on April 8 2020, 91 and 34 days after the first and the last admission, respectively. The file of each patient along his/her hospital course was cautiously reviewed, including whenever the patient was transferred to another hospital. The following data were collected for each patient: age, sex, date of admission and discharge in the hospital as well as the vital status at discharge (dead or alive), date of admission and discharge in the ICU as well as the vital status at discharge (dead or alive), beginning and end dates of mechanical invasive or noninvasive ventilation procedures. Whenever a patient was transferred from the ICU in a given hospital to the ICU of another hospital, we considered that such a continuum constituted a single ICU stay.

Since the objective of this study was an assessment of the ALOS in ICU of COVID-19 patients, 8 of the above-mentioned 67 stays were excluded from the analysis: First, one of the 57 patients with a confirmed RT-PCR positive test had contracted COVID-19 at the hospital while this patient was hospitalized for post-complications after a kidney transplantation, and the record file highly suggested an ICU stay relating more to these complications than to COVID-19 infection. Conversely, only three of the ten patients with the radiological evidence of viral pneumonia were included in the analysis: seven patients had clinical characteristics suggesting that the ICU stay might be not mainly related to COVID-19 (e.g. liver lesions, massive cerebral infarction, …), and were therefore excluded from the study.

### Statistical Analyses

Data are expressed as mean (95% confidence interval (CI)) or median [interquartile range (IQR)], and were represented according to the Kaplan-Meier estimator.^15^ In addition, we examined how the ICU ALOS estimates of COVID-19 patients issued from two estimation methods evolve and compare while the cumulative number of available stays increases along the course of the outbreak. All analyses were made with R statistical software version 3.6.1 and censored data were fitted with the use of the *flexsurv* package. The two methods compared were the following:

#### Discharged patients’ estimation (DPE)

This first method applies a straight-forward calculation: all ICU stays of the series for which the discharge date is before or equal to a given follow-up date of interest were considered (and only such stays were considered). Reported ALOS estimate was the mean LOS of those already discharged patients. Reported LOS median and quartiles were calculated on the same patients.

#### Censored patients’ estimation (CPE)

This second method takes into account the inherent censored characteristic of longitudinal data: considering a given follow-up date of estimation, all previously admitted patients were considered, whether or not they were already discharged. A parametric distribution (e.g. exponential, gamma or Weibull) was fitted to the whole set of patients. Such a method for appropriately analysing time-to-event censored data belongs to the standard framework of methods of survival analysis.^15,16^ Reported ALOS estimates, as well as LOS medians and quartiles, are predictions based on this parametric model.

#### Additional comparisons

In order to demonstrate the generalizability of our results, these two methods were also compared using two simulation studies. Both considered a 30-bed ICU with as many patients admitted on day 0 and new patients admitted as soon as the previous ones were discharged. In the first study, simulated LOS were sampled with replacement from the 59 observed LOS in ZHWU. Such a simulation allows to be free from the observed schedule in practice, including the order of occurrence of the lengths of stay observed. The simulation forces the ICU to be initiated in an already saturated functioning admitting COVID-19 patients. The LOS of the patient still in the ICU at the date of last follow-up was imputed. In the second study, LOS were sampled from a parametric gamma distribution in order to explore how estimates evolve with time in a situation where the true distribution is known.

## 3. Results

The median age of the patients was 62 years old [IQR 52–70] and 38 (64%) were men. The time-course of the ICU stays of the 59 COVID-19 patients is shown in figure 1A. At the date of last follow-up, April 16, one patient was still in the ICU, 21 deaths (36%) had occurred in the ICU, and the 37 patients discharged alive from the ICU were also all discharged alive from the hospital. Invasive mechanical ventilation procedures concerned 40 (68%) patients: stays involving only noninvasive ventilation concerned 11 patients, stays involving only mechanical invasive ventilation concerned 12 patients, and 17 patients had shifted from one type of ventilation to another during the course of their stay. The mean and median estimates for the duration of mechanical invasive ventilation was 21.6 days (95% CI 15.4–28.7) and 12.0 days [IQR 8.5–31 days], respectively. The corresponding estimates for noninvasive ventilation were 5.6 days (95% CI 3.9–7.8) and 3.5 [IQR 1.9–3.0], respectively.

**Figure 1.**
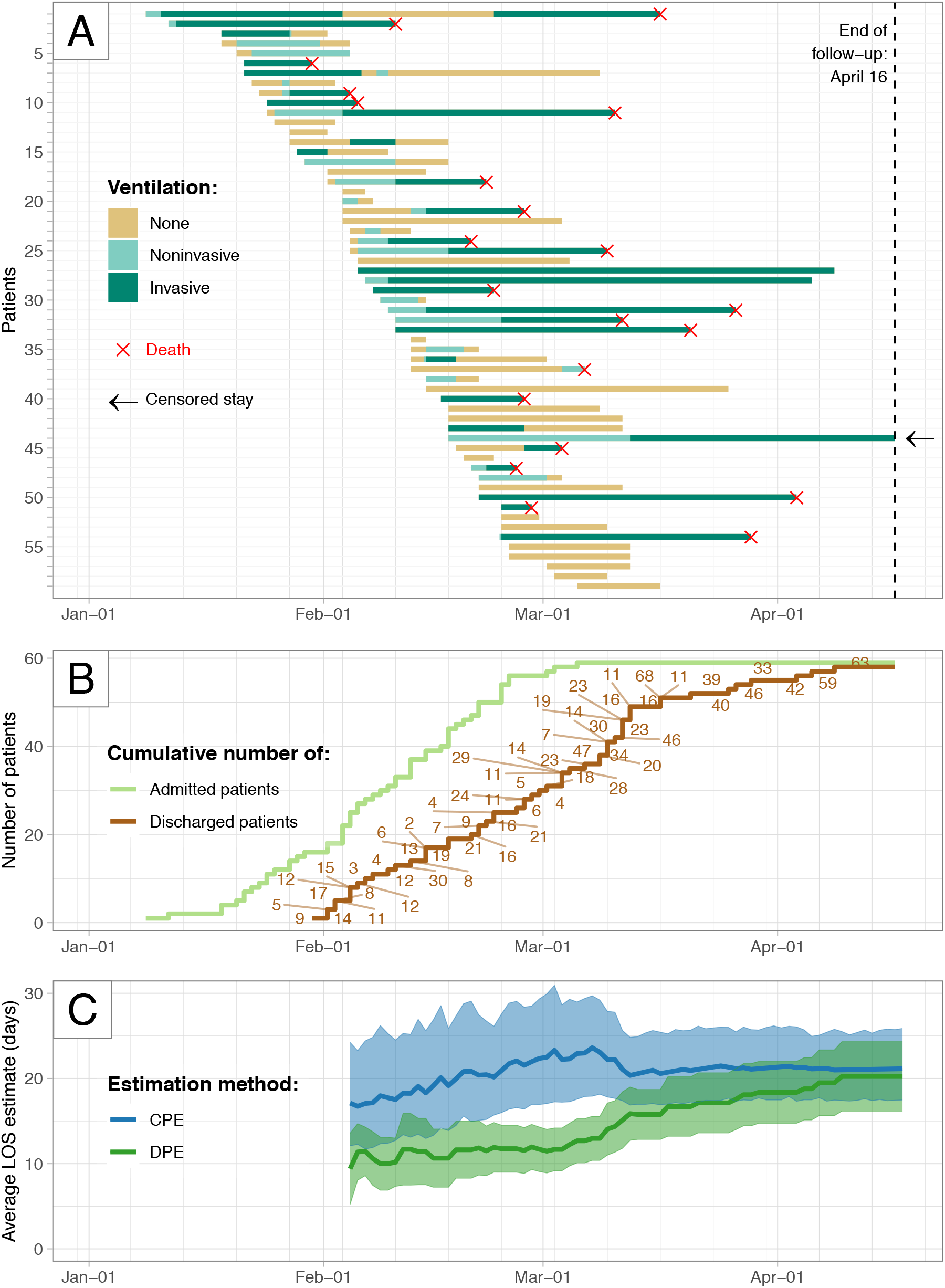
Timeline of the 59 COVID-19 cases treated in the intensive care unit (ICU). Panel A: For each case are plotted admission and discharge dates, periods of ventilation, and vital status at discharge. Panel B: Cumulative numbers of admission and discharge according to time. Panel C: Evolution of the estimates of ICU average length of stay (LOS) issued from the two methods of estimation according to the date chosen for estimation. The expected estimate is shown together with the corresponding 95% confidence interval. CPE, method including censored cases; DPE, method considering only stays for which the patient was already discharged from ICU at the date of estimation.

Figure 1B shows the cumulative number of admissions and discharges according to time. At the date of last follow-up, over 3 months (99 days) had passed since the date of the first admission, January 8. Figure 1C shows the evolution of DPE and CPE-based ALOS estimates according to the accumulating data that become available as time passes. Exponential, Weibull, and gamma distributions led to similar fits of the data – with a delayed convergence for the exponential distribution – and we retained the gamma distribution for reporting CPE. On February 8, one month after the first admission date, 11 (19%) patients had been discharged from the ICU and the corresponding estimate of ALOS with DPE was 18.0 days (95% CI 12.2–27.0), nearly twice that of DPE which was 10.0 days (95% CI 6.9–13.1). On March 8, two months after the first admission date later, 38 (64%) patients had been discharged and the estimate of ALOS issued from DPE and CPE at that date was 13.0 days (95% CI 10.4–15.6) and 23.1 days (95% CI 18.1–29.7), respectively. Under the assumption that the parametric distribution used in CPE is well fitted to the data, the two methods should converge towards a similar estimate when no censoring data remain, *i*.*e*., when all patients of the cohort have been discharged: while only one patient remained in the ICU at the last date of follow-up, April 16, ALOS with CPE and DPE was 21.1 days (95% CI 17.5– 25.3) (which constitutes the most accurate estimate on the study data set) and 20.2 days (95% CI 16.2–24.3), respectively. CPE estimate of hospital ALOS 30.6 days (95% CI 26.2–35.3) and median duration of hospital LOS was 27.0 days [IQR 16.5–39.0]. Whenever some patients of the cohort remain treated in the ICU at the date of follow-up, DPE yields a biased underestimation of ALOS: discharges observed early are more likely to concern patients with a short LOS or conversely, the discharges occurring at the end of the process are more likely to concern patients with a long LOS. Fig 1B, in which the LOS corresponding to each discharged patient is indicated along the discharge curve, illustrates this pattern: nine out of the 10 first occurring discharges concern LOS < 15 days, while eight out of the 10 last occurring discharges concerned LOS > 30 days. In the end, the simulations shown in the Supplementary Appendix 1 demonstrate the generalizability of the biased pattern of DPE, and the unbiased pattern of CPE. Figure 2A presents a Kaplan-Meier estimator and indicates that the median ICU LOS is around 16 days. The corresponding estimate issued from CPE is slightly higher, at 17.4 days, because the corresponding parametric fit is impacted by the substantial frequency of very long stays: Figure 2B shows the LOS distribution and 14 out of the 59 patients (24%) had a length of stay ≥ 30 days. The relatively high frequency of such patients with a very long LOS explains why the expected estimates of ALOS shown in Figure 1C requires a substantial delay until remaining stable. Interestingly, among the 14 patients with a LOS ≥ 30 days, 8 had died while the total number of observed deaths in the cohort was 21. The fact that 38% (8/21) of the deaths observed occurred in patients who had an ICU stay ≥ 30 days also indicates that obtaining a reliable estimate of the mortality rate in the patients admitted to the ICU as well as obtaining a reliable ALOS of the individuals dying in the ICU also requires waiting a substantial delay after the beginning of the outbreak.

**Figure 2.**
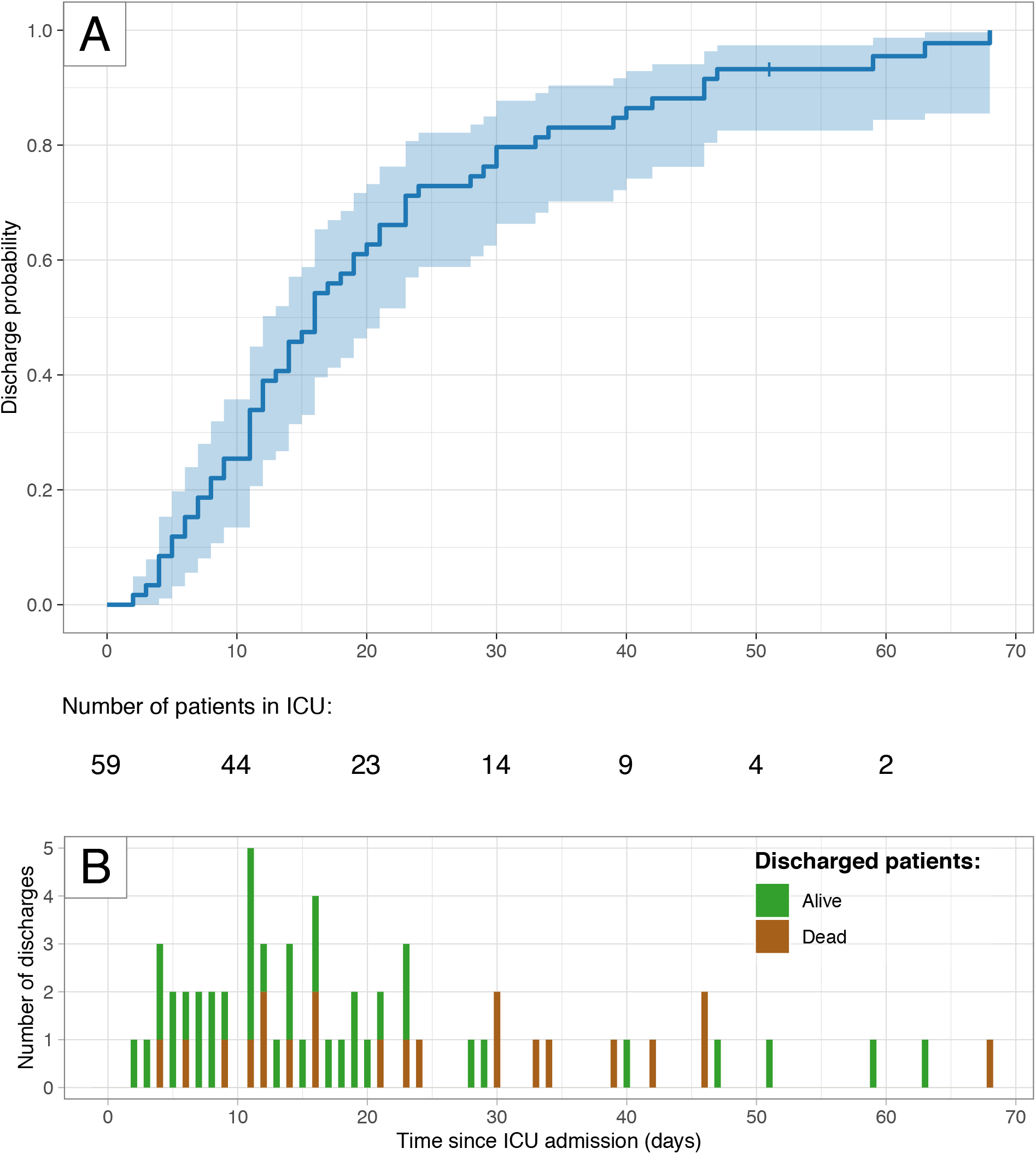
Discharge data according to time since intensive care unit (ICU) admission. Panel A: Kaplan-Meier representation of the probability of discharge according to ICU length of stay (LOS). Panel B: Distribution of the observed ICU LOS.

## 4. Discussion

Taking the COVID-19 outbreak as an emblematic example of the first outbreak of a threatening pandemic due to a novel infectious agent, the present study demonstrates the importance of obtaining a reliable estimate of the ICU ALOS in such situations. The study also recalls that appropriate methods of estimation require the inclusion of censored cases in the analysis, and we also demonstrate the important bias associated with calculations only based on the stays of already-discharged patients. Finally, whenever patients of the population treated in the ICU with a long LOS are observed at a substantial frequency, as was observed in the present reported series, the bias relating to inappropriate methods might be especially important.

Although the present study shows that ALOS constitutes an important parameter, we failed to find any observational study of COVID-19 cases to date that reported ALOS. Nevertheless, several studies to date have reported median estimates of ICU LOS and such a choice is perfectly understandable: since ICU LOS is not normally distributed, a reporting of median and IQR instead of the mean is recommended. The median of ICU LOS reported to date^6-8,17,18^ (see Table S2 in the Supplementary Appendix 2) often concern a particular sub-population (e.g. patients who died, patients who survived), ranged from 4 days (estimate considering six patients who died in the ICU) to 11 days (estimate reported in the same study and based considering 12 patients discharged alive from the ICU)^17^, and raise concerns in terms of the potential bias of the reported estimates (see Table S2 in the Supplementary Appendix 2). These concerns may be then extended to modelling studies^9,10,12,13,17^ that will naturally parameterize their forecasts according to the observational data reported (Table S2 in the Supplementary Appendix 2). The data of the series reported here yielded an estimate of ICU ALOS at 21.1 days (95% CI 17.5–25.3) and a median ICU LOS at 17.4 days [IQR 9.6– 28.7]. These estimates are well above estimates reported to date. These estimates are associated with several strengths. First the whole study time course lasted 99 days, enough time had passed for allowing a last date of follow-up at which all patients but one were discharged. To our knowledge, such a resulting quasi complete distribution of the LOS observed in a given series of COVID-19 cases (see Figure 2B) has not been reported to date, and in addition, such a data set is indeed appropriate for assessing estimation methods since the target value of the estimate is nearly perfectly known (only one stay remained censored). Second, the high values reported here are based on a reasonable sample size (n = 59) and our study demonstrates that an unbiased estimate at a reasonable distance from the beginning of the epidemics is inherently higher than that issued from a biased calculation a short time after the beginning of the outbreak. Nevertheless, our study also has some limitations. The study is monocentric and therefore, the extrapolation of our estimates to other settings is questionable. Moreover, cultural behaviors, modifications of triage decisions, changes in the management of patients may vary not only from one place to another, but also with time according to the pressure of this threatening epidemics on the organization of healthcare workers. However identical drawbacks would also stand for most studies reported to date. The main outcome of this study is alerting the community about three elements. First, all scientists working on COVID-19 must realize that when dealing with data relating to LOS, they should imperatively use appropriate methods devoted to the analysis of censored data. Such methods are not original, they belong to the standard tools used in the domain of survival analysis and are easily available in any statistical software. There is no reason for avoiding their usage, and the reader will find an illustrative computer code in the Supplementary Appendix 3. An additional strength of these methods – not explored in this article but illustrated in Supplementary Appendix 3 – is their ability to fit individual characteristics of patients with multivariable models to predict LOS in specific strata of the population in order to adapt to varying recruitment settings. A side result of the analyses made in the present study suggests that the death rate of COVID-19 patients in the ICU might also be underestimated, and on this topic, the present study shares many perspectives with the work of Lipsitch et al on the biases associated with the estimation of case-fatality risks.^19^ Second, in the context of the first outbreak of a novel infectious agent, some estimates concerning time-to-event data such as hospital LOS, ICU LOS, duration of ventilation, time of illness onset to ICU admission, etc. constitute a kind of critical food required to feed forecast models and these models are very important in many issues such as exploring and comparing mitigation scenarios, or optimizing preparedness. Therefore, enhancing the quality of the above-mentioned estimates is an important concern and our study suggests that there is room for such enhancements in the analyses of COVID-19 epidemic. Third and to conclude, whenever the estimates reported in this study would be generalizable to other settings, then this is bad news: long ICU LOS as reported here imply that occupied beds remain unavailable for a long time and this adds additional pressure to the surge in ICU beds encountered in many places worldwide.

## Data Availability

Data are not available.

## 7. Author Contribution

The decision of performing the study emerged from informal discussions involving NL, XZ, FC, BR, YZ, and GH. Study conception and design: NL and GH. Data acquisition: XZ and YZ had full access to all of the raw data in the study and can take responsibility for the integrity of the data. Analysis: NL and GH. Interpretation of data: NL, XZ, FC, BR, YZ, and GH. First draft of the article: NL and GH. All authors approved the final version of the article. Authors declare that they have no competing interests.

